# Automating Handwritten Vaccination Record Transcription with Generative Multimodal AI Models: A Proof of Concept Study from The Gambia

**DOI:** 10.1101/2025.07.08.25329577

**Authors:** Roy Burstein, Alieu Sowe, M. Carolina Danovaro-Holliday, Mitsuki Koh, Joshua L. Proctor

## Abstract

**Background:** Handwritten home-based vaccination records (HBRs) are a vital source of immunization data, yet manual transcription in household surveys is time-consuming, error-prone, and resource-intensive. Recent advances in multimodal generative AI models offer a potential pathway to partially automate this task under certain conditions, improving efficiency while maintaining data quality.

**Methods:** We evaluated the performance of generative multimodal AI models, specifically OpenAI’s GPT-4o, in transcribing handwritten vaccination cards from a 2022 survey targeting children aged 12 - 35 months in The Gambia. Using a curated dataset of 335 cards (6,700 vaccination entries), we developed a gold-standard benchmark from three human transcribers (all of them in The Gambia) and assessed AI model performance across transcription accuracy, vaccination coverage estimates, timeliness, and missed opportunities for simultaneous vaccination (MOSV). We also tested a confidence-based segmentation approach to identify high-confidence transcriptions suitable for automation versus low-confidence entries requiring human review.

**Results:** The fine-tuned GPT-4o model achieved 79% accuracy for exact date transcription and reached human-level performance (94% accuracy) on 69% of entries classified by the AI as high-confidence. Coverage and timeliness estimates from high-confidence transcriptions were 98.0% and 91.6% accurate, respectively, compared to 98.8% and 95.6% from human transcribers. Date errors by AI and humans differ systematically, with AI showing fewer year-shift errors.

**Conclusion:** Multimodal AI models show strong potential for automating HBR transcription in immunization coverage surveys, at least in a setting like The Gambia. When paired with confidence-based filtering, these models achieve human-level performance on coverage and timeliness estimates—the key metrics used in programmatic decision-making—across a large subset of records. This enables substantial gains in efficiency while preserving data quality. Further research should evaluate generalizability across diverse card formats, languages, and contexts to support integration into real-world immunization programs and health monitoring activities.

## Introduction

The transcription of handwritten home-based vaccination records (HBRs) is a central component of immunization coverage monitoring through household surveys (1). In addition to caregiver recall, which is a less reliable source of information (2), these records serve as primary sources of immunization history in household surveys (3) and are the foundation for estimating key indicators of immunization program performance, such as vaccination coverage, numbers of children unvaccinated, timeliness of vaccination (4), and missed opportunities for vaccinations during healthcare visits (5). Such indicators are regularly monitored by ministries of health, as well as global bodies such as the World Health Organization (WHO) and the United Nations International Children’s Emergency Fund (UNICEF) (6), modelling groups such as the Institute for Health Metrics and Evaluation (IHME) (7,8), WorldPop, and other supporting disease-specific initiatives (21,22), and serve as benchmarks for global goals such as the Sustainable Development Goals (SDGs) (9) and the Immunization Agenda 2030 (IA2030) (10). Beyond monitoring, these indicators also play a critical role in intervention planning, guiding adjustments to routine immunization systems and providing evidence for the need for supplementary immunization activities to address gaps in coverage (11,21,22).

Globally, the number of vaccination cards transcribed annually is substantial, spanning large-scale national surveys such as the Demographic and Health Surveys (DHS), Multiple Indicator Cluster Surveys (MICS), and other immunization coverage studies, including many post-campaign coverage surveys and related assessments (23). The WHO Vaccination Coverage Cluster Survey Reference Manual provides standardized guidance for field-based transcription, where data collectors manually record information from vaccination cards during household visits. Pictures of HBRs are often taken, but they are mainly used to verify data when inconsistencies are identified. However, despite their importance and broad use, manual transcription of HBR presents significant logistical challenges. It is a time-intensive task, particularly in large-scale surveys, and is prone to human error, which could introduce biases in coverage estimates, and other survey indicators such as timeliness and vaccination drop-out (24,25,30). In a recent study conducted in The Gambia, field-based transcription took an average of six minutes per card, contributing significantly to survey implementation time and costs (26). At this rate, a study reviewing 20,000 cards would take about 1-full time employee-year’s worth of labor. Also, interviewer fatigue may reduce requests cards for transcription (27). In addition to saving time, there is the opportunity to decrease the rate of errors inherent in survey data collection (20).

Given these challenges, there is interest in improving the typical field transcription workflow. For example, by taking images of HBRs in the field and doing transcription later based on these images, a workflow has been used in a few settings (28), but rarely documented. This approach was tested, vis-a-vis the usual workflow of card transcription by surveyors in the field, in a survey study from the Gambia, where image-based transcribers were able to accurately transcribe cards in about half the time it took in the field. Image-based transcription has several potential advantages, such as saving time in the field, and can potentially reduce errors by utilizing specialized transcription experts, conducting duplicate transcriptions, and maintenance of original records for further review and provenance.

Generative artificial intelligence (AI) models can be leveraged to augment image-based transcriptions by humans. These multimodal models, such as OpenAI’s GPT-4o, integrate both visual and language processing capabilities, allowing them to interpret handwritten text, recognize structured form layouts, and extract meaningful information directly from images (12). Unlike traditional computer vision algorithms that rely on convolutional neural networks (CNNs) or other optical character recognition (OCR) techniques, transformer-based neural networks, in particular vision transformers (ViTs) enable a more advanced approach by analyzing images in a way that captures long-range, spatial relationships and contextual cues (13); recent reviews comparing ViTs and CNNs highlight how ViTs have better performance for applications in medical image analysis (14). The addition of large language models (LLMs) further enhances this process by providing reasoning and contextual interpretation of extracted text, an area where conventional OCR methods often fall short (15). This combination has the potential to improve transcription efficiency and accuracy while minimizing reliance on manual data entry. Furthermore, AI model transcription failures may be predictable, enabling targeted review processes that enhance overall accuracy and reliability.

In this study, we explore the feasibility of using these state-of-the-art, generative multimodal AI models to transcribe vaccination cards from images with high accuracy. Here, we leverage a curated benchmark dataset for validation to rigorously assess transcription accuracy. Specifically, we analyze the accuracy of transcriptions for exact dates on handwritten records, in addition to estimating key immunization performance indicators. We also implement an approach to quantifying model uncertainty, in order to identify transcription failures for human review. By evaluating the accuracy of AI-assisted transcription and its implications for immunization data collection, we aim to provide insights into how AI can be integrated into realt-time, coverage monitoring workflows to enhance efficiency and accuracy for global vaccination programs.

## Data and Methods

For this study, we reanalyze data from an operational study conducted in The Gambia in 2022 to assess different approaches to transcribing vaccination data from HBRs (5). The study compared field-based transcription, where data collectors manually recorded vaccination dates from cards during household visits, to image-based transcription, where HBRs were photographed by the surveyors, using the same computer-assisted personal interviewing (CAPI) tool, CSPro, used for the survey questionnaires, and later transcribed by a centralized team. This produced a dataset consisting of 349 cards, each of which was associated with transcriptions performed by three human transcribers: the field surveyor and two individuals in an office in Banjul. Fieldwork took place in September 2022, and central data extraction in September and October 2022. The official vaccination card in The Gambia at the time had space to record BCG, three doses of pentavalent vaccine (diphtheria, tetanus, pertussis, hepatitis B and *Haemophilus influenzae* type b), oral polio vaccine birth dose (or polio 0) and five other OPV doses, one dose of inactivated polio vaccine (IPV), three doses of pneumococcal conjugate vaccine (PCV), two doses of rotavirus, one dose of yellow fever, one dose of meningitis A, and two doses of a measles-rubella vaccine. Of relevance to the findings, the meningitis A vaccine was introduced in February 2019 and some old card models did not include space for this vaccine. Dates on HBRs were all hand written, including various idiosyncrasies such as writing in margins, writing the same date across multiple boxes, and using hatch-marks to likely indicate ‘same as above’. Furthermore, there were at least three different card designs in the data, owing to different ministry of health versions, some from private facilities, or children coming from other countries (Figure 1).

**Figure 1:**
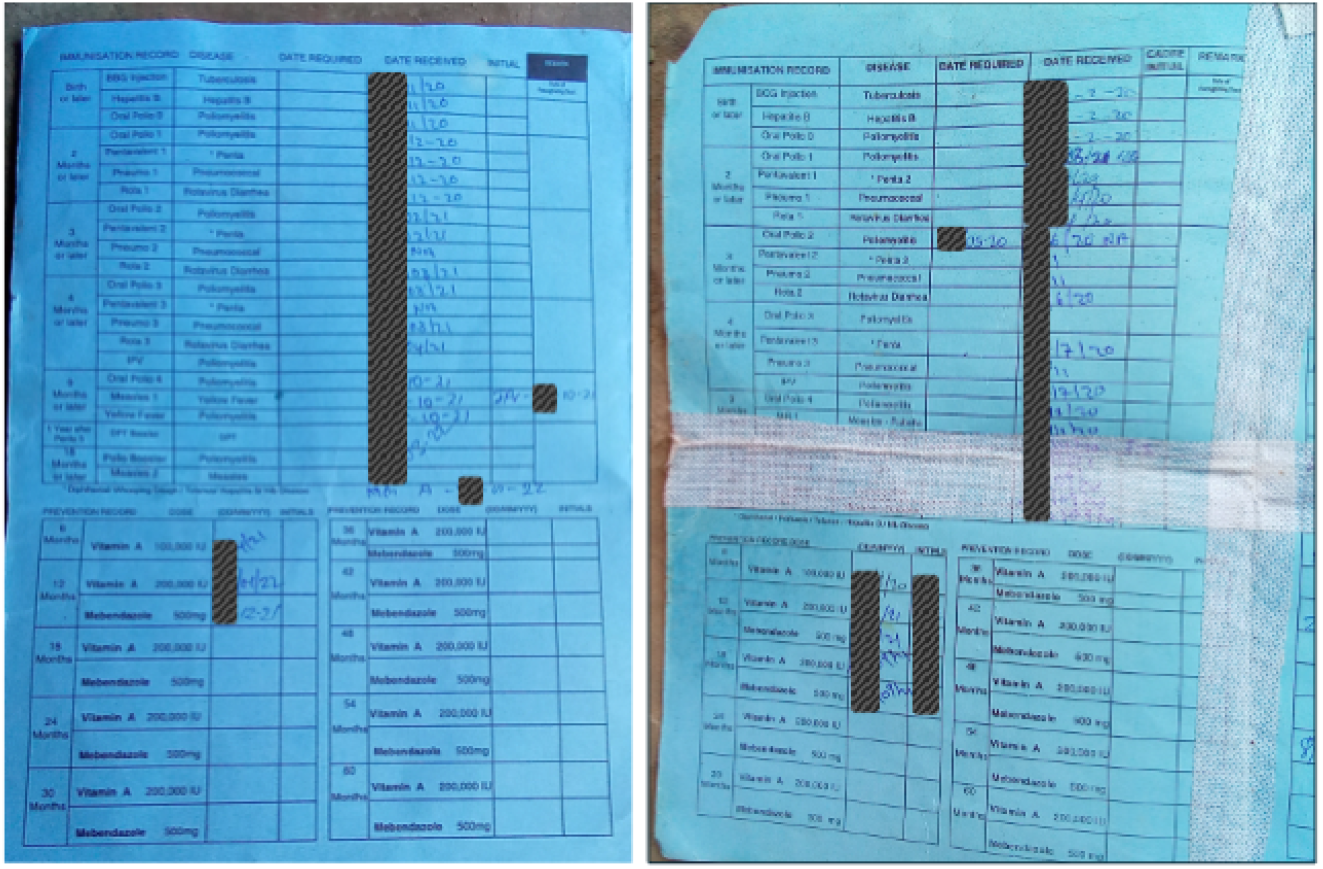
Two example vaccination records used to demonstrate digitization complexity for both manual and AI transcription. Identifying details, including exact dates, have been visually redacted (dark grey boxes) to ensure privacy.

For the comparison with AI, we performed several preprocessing steps to ensure a standardized and high-quality dataset. We verified that each vaccination card was correctly matched to its corresponding transcriptions. Some cards lacked a complete set of three independent transcriptions, leading to their exclusion. We also reviewed and refined the images, ensuring they were cropped and oriented for optimal readability. Additionally, images that were low-resolution or poorly lit were deemed unusable. After applying these quality control measures, 24 cards were discarded, resulting in a final curated dataset of 335 vaccination cards with corresponding transcriptions. Child birth dates were collected at the time of the survey; however, all card images were scrubbed of personal identifiers during the original data collection.

To establish a gold-standard transcription for each vaccination entry, we first assumed that a transcription was correct if all three human transcribers from the original study agreed. However, in 14% of cases, there was disagreement among the transcribers. In these instances, in 2024, one author (RB) manually adjudicated the correct transcription by reviewing the conflicting entries alongside the corresponding HBR image. Twenty individual vaccination doses per card were tracked, resulting in a dataset of 6700 validated transcriptions.

We used OpenAI models to transcribe vaccination cards, accessed via their API. Initial tests were conducted using GPT-4o-mini and GPT-4o (2024-08-06 version), followed by a fine-tuned version of GPT-4o (2024-08-06 version) for the final analysis. To fine-tune the model, we split the dataset into 168 cards for training and 167 cards for testing. The fine-tuning process involved using card images as input and their corresponding gold-standard transcriptions as output. Our full transcription prompt is available in the project’s code repository.

We assessed exact date transcription accuracy by calculating the proportion of correctly transcribed dates, using the gold-standard transcription as the reference. Additionally, we evaluated transcription error magnitude by measuring the absolute difference in days between the AI-generated transcription and the gold-standard date. For vaccination coverage accuracy, we first converted dates into binary indicators, where the presence of a date indicated that a vaccine dose was administered. This allowed us to compute both overall coverage and coverage accuracy. Timely coverage was defined here using a strict definition, as a vaccine administered within 14 days before or after its scheduled due date, based on the child’s birth date (29). Missed opportunities for simultaneous vaccination (MOSV) were calculated as the proportion of cases where co-administered vaccines, defined as those scheduled for the same visit, were not administered together (30).

To improve the efficiency of human review, we developed methods to distinguish high-confidence transcriptions from those requiring further validation. Identifying low-confidence transcriptions enables implementers to prioritize human resources effectively, focusing efforts where they are most needed.To accomplish this, we tested multiple approaches to segment transcriptions based on predictive confidence metrics relative to the gold standard. One approach involved prompting the model to self-assess transcription accuracy, sometimes called self-reflection, however this method ultimately proved unreliable (see Supplementary Information). We found that using multiple candidate transcriptions (model draws) per card image, varying the model temperature, and analyzing the consistency of the output across draws provided a more effective way to predict accuracy (16). By observing multiple generated completions from the model, this approach draws insight from the model’s inherent uncertainty distribution, even without access to the underlying LLM or tokenization model. For this study specifically, if all candidate transcriptions for a given card agreed, the transcription was classified as high confidence. To further refine this approach, we tested different combinations of model temperature and number of draws to explore trade-offs between the size of the high-confidence segment and its overall accuracy. This allowed us to balance maximizing the proportion of data classified as high confidence while, importantly, maintaining a level of accuracy comparable to human transcribers.

All code for this analysis is available at https://github.com/rburstein-IDM/ai_vxcard_transcription

### Data availability statement

The data supporting this study’s findings are available upon submission of a reasonable request and a data-sharing agreement.

### Ethics review statement

This study utilised data from an operational study conducted in The Gambia. That study was approved by the Scientific Coordinating Committee of the Medical Research Council Unit The Gambia at the London School of Hygiene and Tropical Medicine and The Gambia Government/MRCG Joint Ethics Committee (Project ID/Ethics ref: 27860). The present study was cleared by the University of The Gambia Research Ethics Committee (UTGREC) (Ethics ref:UTG-REC/09/2024/0047).

## Results

Across 335 vaccination cards, each tracking a maximum of 20 vaccine doses, the dataset comprised 6,700 validated transcriptions. After splitting the data for fine-tuning, the test set consisted of 167 cards and 3,340 transcriptions. Among the three human transcribers from The Gambia, 86.0% of transcriptions were in agreement, and the overall accuracy compared to the adjudicated gold-standard transcription was 94.0%. The field-based transcriber achieved 91.7% accuracy, while the two image-based transcribers achieved 94.4% and 95.9%, respectively. AI-based transcription performance improved across model versions, with overall accuracy for exact date transcription at 47% for GPT-4o-mini, 63% for GPT-4o, and 79% for the fine-tuned GPT-4o.

Varying the model parameter temperature and multiple draws from the model with the same input allowed for the segmentation between high and low confidence transcription images (*Figure 2*). At one extreme, using a single draw in consensus includes all transcriptions, but at a lower accuracy of 79%. At the other extreme, requiring full consensus across draws achieves 97.5% accuracy, but only 50% of the data qualifies for the high-confidence segment. Between these extremes, the trade-off follows a roughly linear pattern, where increasing the number of draws improves accuracy but reduces data retention. The efficiency of this trade-off improves at higher temperatures, allowing us to reach a 94% human-level accuracy benchmark with fewer draws. Specifically, this accuracy is achieved at: 5 draws at temperature = 1.3; 10 draws at temperature = 1.0; over 30 draws at temperature = 0.7; and not yet achieved by 40 draws at temperature = 0.4. In each case, once the 94% accuracy threshold is met, approximately 69% of transcriptions qualify for the high-confidence segment. Based on these findings, for the remainder of our results, we use temperature = 1.0 with 10 draws in consensus as our segmentation criteria.

**Figure 2:**
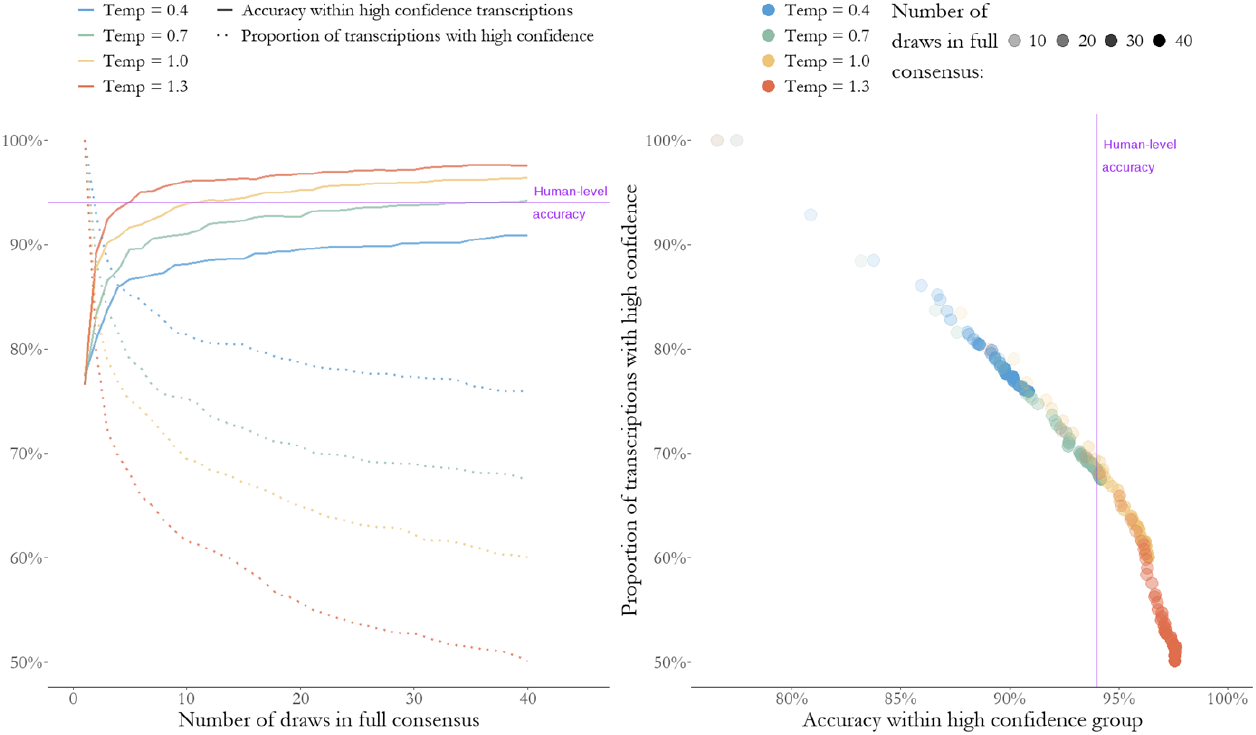
Trade-offs between accuracy and data inclusion in model confidence segmentation. The purple line represents human-level accuracy (94%), showing where different configurations achieve this benchmark. Left: The relationship between the number of draws in full consensus and both accuracy (solid lines) and proportion of transcriptions classified as high-confidence (dotted lines), across different temperature settings. Right: The accuracy of transcriptions within the high-confidence segment as a function of the number of draws in full consensus and temperature, with marker transparency indicating the number of draws required and color representing temperature.

It is also notable that this segmentation was reflected by the human transcribers, though to a lesser extent. For instance, human accuracy for the entries labeled high confidence was 95.6%, while among low confidence transcriptions it was 90.6%. The split for the AI was 93.9% and 44.0%, respectively. (*Figure 3*)

**Figure 3:**
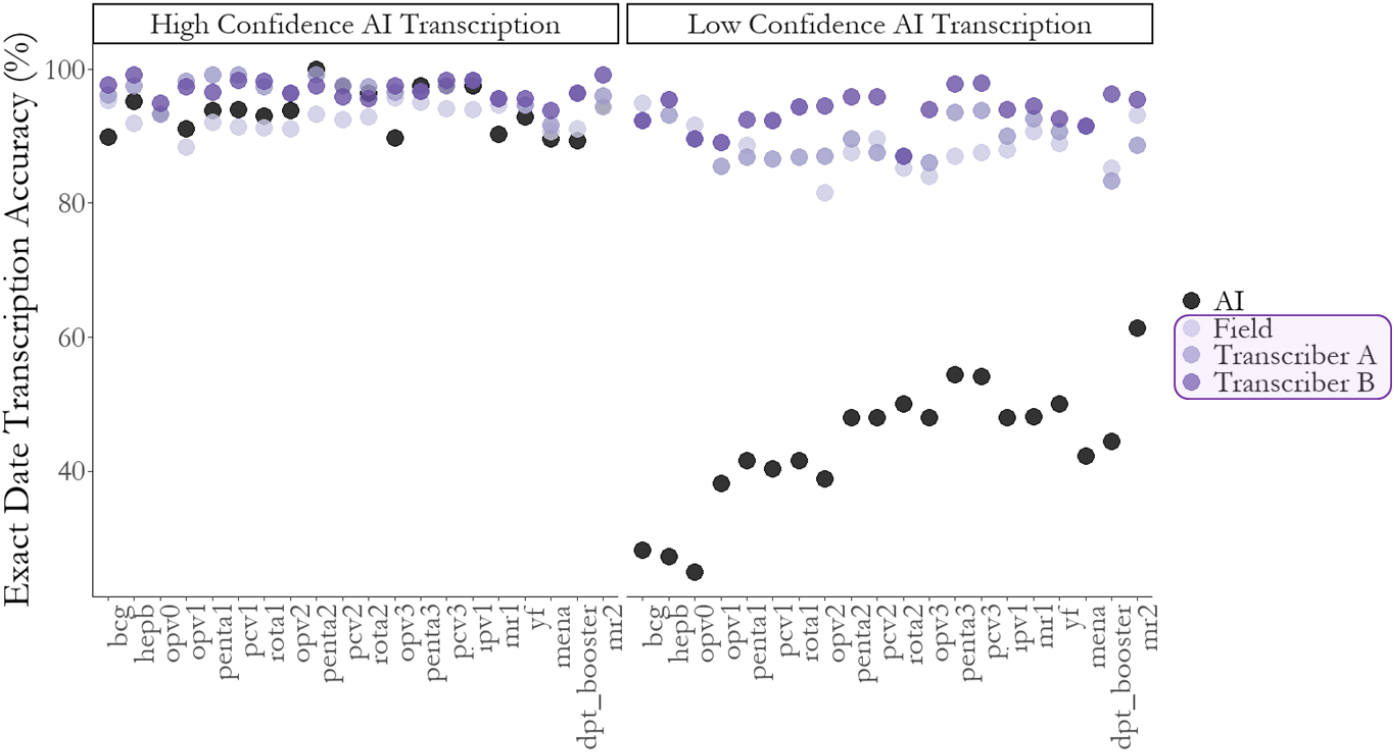
Exact date transcription accuracy by vaccine, AI and three human transcribers compared to gold standard. Left: Among entries segmented as High Confidence. Right: Among entries segmented as low confidence.

Across the 20 vaccines, the proportion of transcriptions classified as high confidence typically ranged between 67% and 77%, with one notable exception: Meningitis A, where only 57% of transcriptions met the high-confidence threshold. This lower confidence rate was likely due to its placement on the vaccination card, as it often appeared along the fold, hampering legibility. Additionally, some card designs omitted Meningitis A, leading to cases where it was handwritten in the margins, further reducing transcription consistency and legibility.

Among incorrectly transcribed dates, 73% of those classified as high confidence and 75% of those in the low confidence segment were accurate within ±30 days. Within ±6 months, 94% and 96% of transcriptions were correct, respectively. These error magnitudes outperformed human transcribers, with the most accurate human achieving only 67% accuracy within 30 days and 86% within 6 months. Additionally, the pattern of errors differed between AI and human transcribers (Figure 4). AI transcriptions, particularly low-confidence outputs, were often off by one month, visible as spikes along the blue lines. Notably, one of the image-based human transcribers frequently made errors exactly one month off (for example writing 22/07/2022 instead of 22/06/2022), suggesting a consistent misinterpretation. In contrast, all three human transcribers were commonly off by an entire year (for example writing 22/06/2021 instead of 22/06/2022), an error that was rare in AI-generated transcriptions. This distinction highlights how AI models and human transcribers introduce different types of systematic biases, which may have implications for testing and integrating AI-assisted workflows in vaccination data transcription.

**Figure 4:**
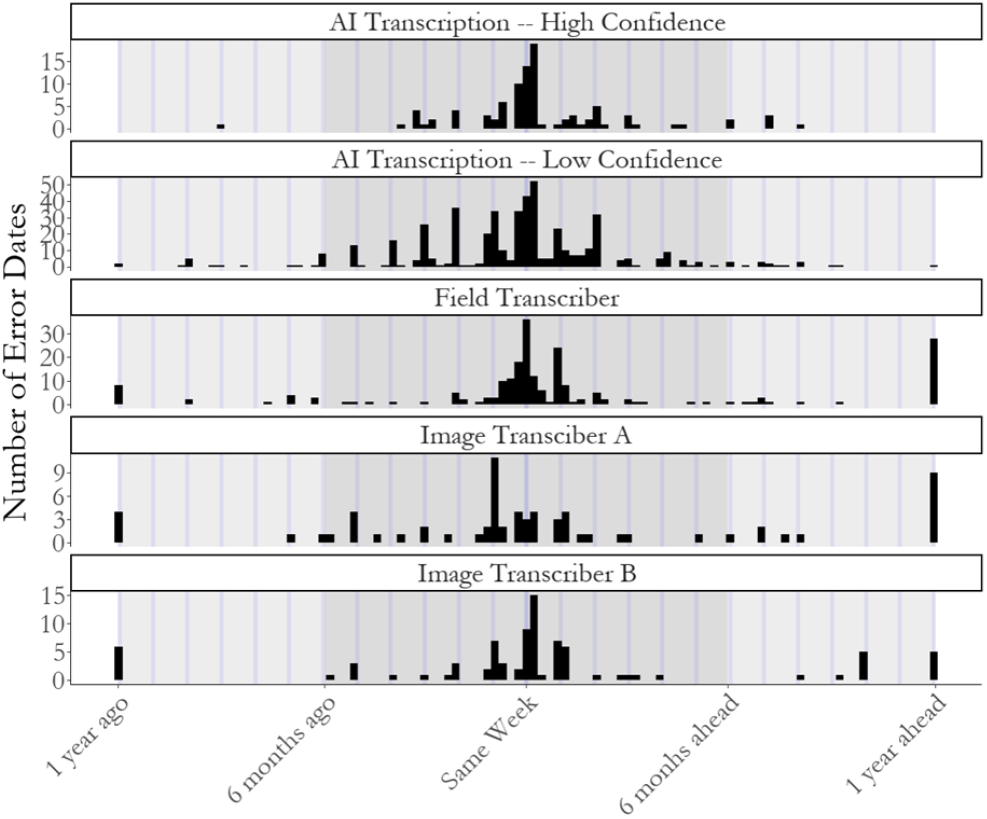
Patterns in date errors among incorrect transcription by AI Confidence segment and 3 human transcribers. Blue lines represent one-month increments from the validated date. Note the higher overall number of errors in the low confidence AI set.

Coverage estimates across all vaccinations were 98.0% accurate among high confidence AI transcriptions, and 91.6% among low confidence AI transcriptions, with average human transcribers achieving 98.8% and 95.6%. Overall vaccination coverage in this sample from The Gambia was high, with all vaccines except for four over 91% *(Figure 5)*. Timely coverage was 99.3% accurate for high confidence AI transcriptions, similar to human level, and 82.7% for low confidence transcriptions. Prevalence of MOSV was frequently underestimated by AI in the high confidence transcription set for each childhood vaccination visit, meaning the dates were more often transcribed as the same as co-administered vaccines, when according to the validated transcriptions they were not *(Figure 6)*.

**Figure 5:**
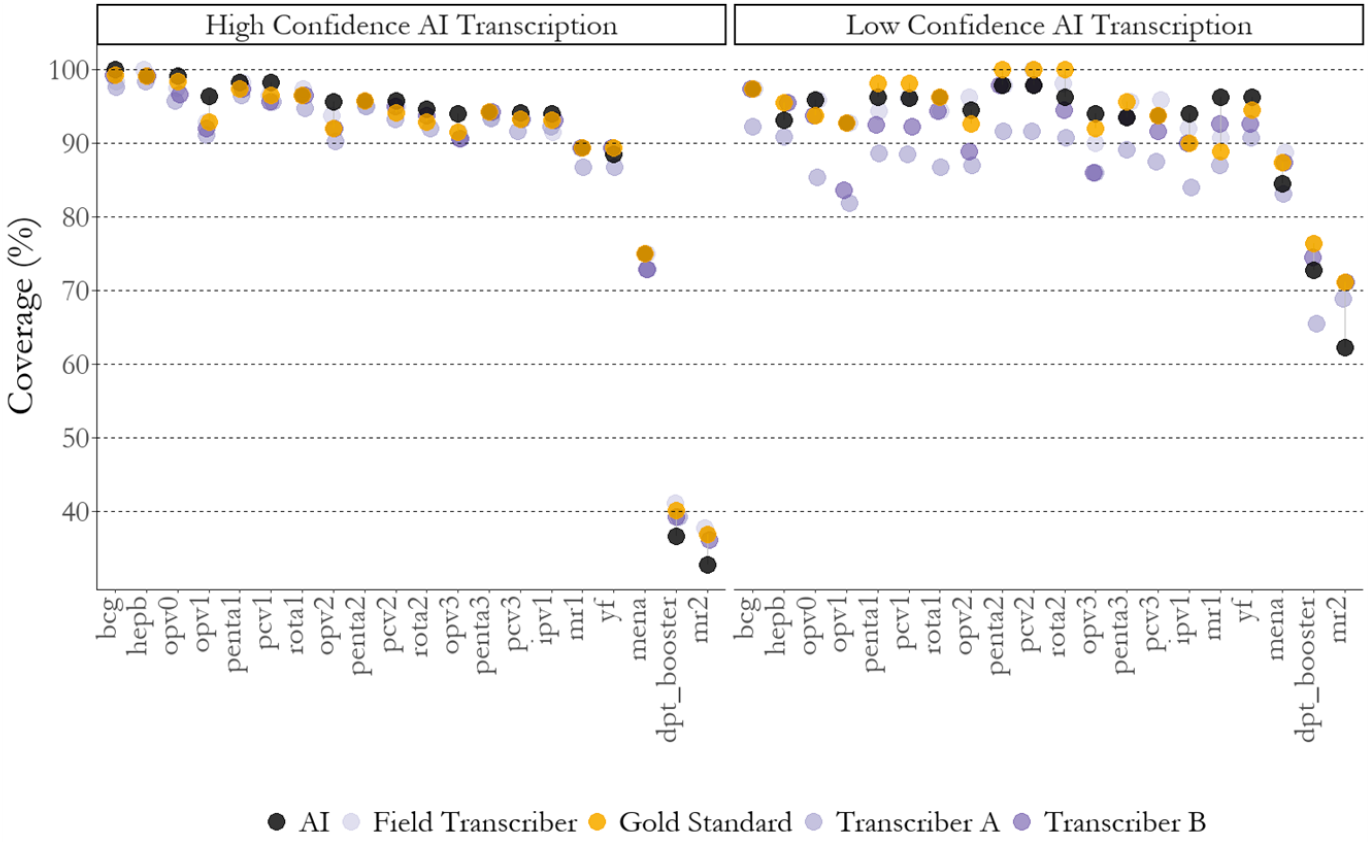
Estimated coverage by gold standard versus different transcribers among high confidence subset (left), and low confidence subset (right).

**Figure 6:**
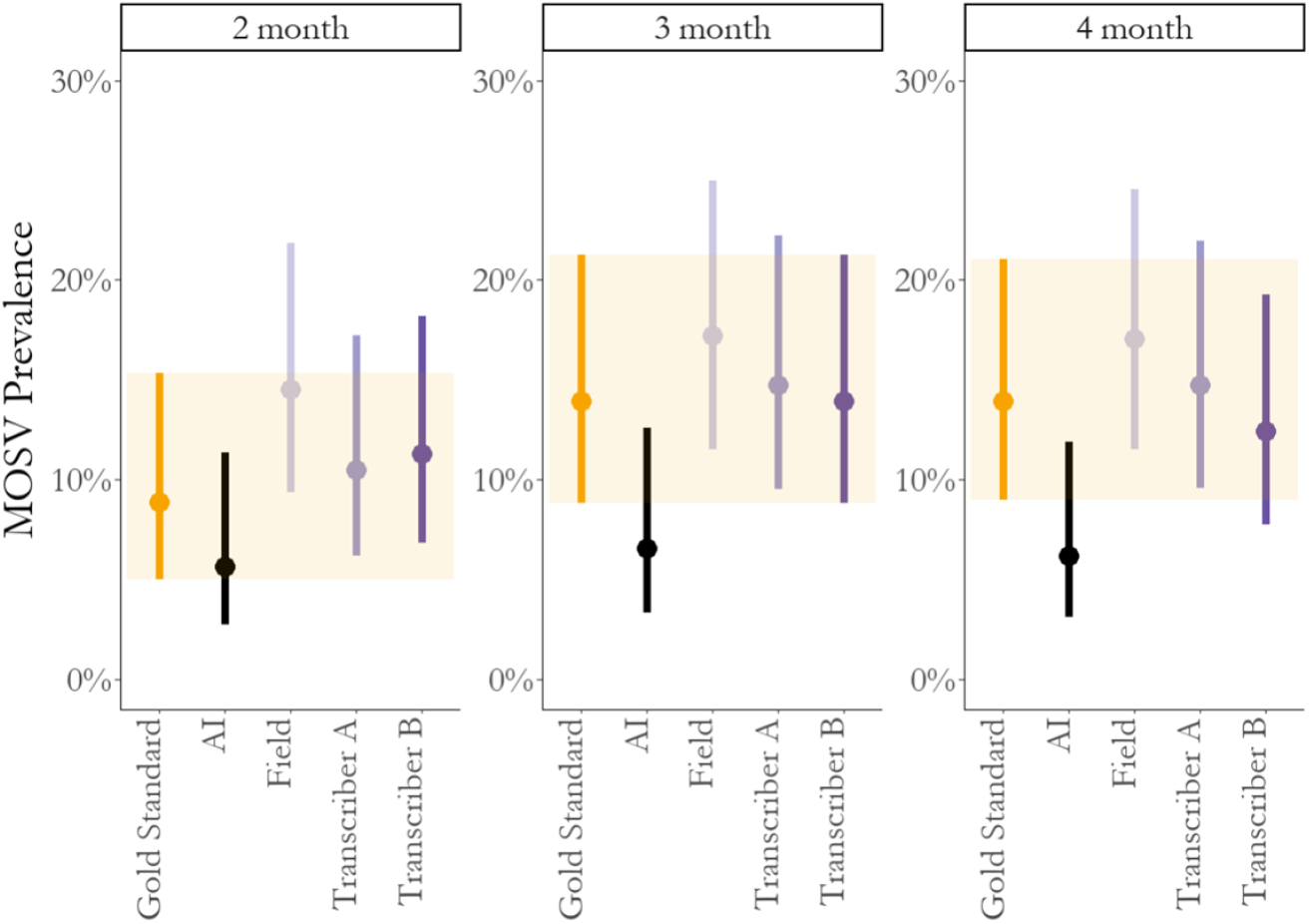
Estimated prevalence of missed opportunity for simultaneous vaccination among the high confidence set of transcriptions by visit, as estimated based on AI transcription versus the three human transcribers, compared to the gold standard estimate.

## Discussion

This proof-of-concept study demonstrates that state-of-the-art (SOTA) multimodal AI models can accurately transcribe complex, handwritten vaccination records from images. Although exact date transcription remains challenging, the models achieved performance comparable to human transcribers in estimating key immunization indicators, such as vaccination coverage (98.0% and 91.6%, AI accuracy of high and low confidence images, respectively, compared to 98.8% and 95.5% for human accuracy) and timeliness. Most date transcription errors were minor, typically falling within one month of the actual vaccination date. Notably, the ability to recover coverage indicators with high fidelity is particularly promising, as these metrics are among the most critical for guiding evidence-based programmatic decisions. We demonstrate an approach for predicting transcription failures, highlighting the utility of confidence-based segmentation to support targeted human review. This approach enhances the scalability and operational feasibility of AI-assisted digitization workflows, with potential applications extending beyond vaccination records.

Through the iterative development of our transcription approach, we identified several key lessons relevant to the integration of multimodal AI-based transcription into future data and research workflows. First, image quality impacts transcription accuracy: proper orientation and removal of non-essential clutter in images improves results. This suggests that either a pre-processing step will be needed to standardize images before AI transcription or that implementers should follow a protocol to ensure consistent and high-quality image capture. Second, model choice matters: we observed a substantial accuracy boost when transitioning from GPT-4o-mini at 47% to GPT-4o at 63%, indicating that SOTA models currently provide the best performance by a significant margin. Finally, fine-tuning models with example images and their transcription further improved accuracy across metrics (79%), suggesting that future implementations will benefit from developing small, domain-specific training datasets to enhance performance.

The generative AI approach for exact date transcriptions across all images achieves moderate accuracy even with fine-tuning of the models. However, we have also developed a novel workflow to identify low-confidence transcriptions and flag them for human review; we believe this approach is essential for practical deployment in real-world immunization data collection workflows. By leveraging consensus across multiple model outputs for the same image, we are able to isolate a high-confidence subset that met or exceeded human transcription accuracy (94% AI accuracy for 69% of the records), while selectively routing lower-confidence cases for manual verification. This approach has significant operational implications. For example, in large-scale vaccine coverage surveys, where thousands or more vaccination cards must be transcribed, automating the majority of transcriptions while minimizing human verification to only uncertain cases can improve efficiency, reduce workload, and maintain data quality. However, this method does increase computational costs due to additional token generation. One way to mitigate this is by adjusting the model’s temperature, which can optimize the balance between accuracy and efficiency. We also expect that the process of human review for difficult to transcribe images will contribute to future iterations of the fine-tuned model ultimately improving both the automatic transcription for exact dates and decreasing the number of manual transcriptions required.

A key advantage of multimodal AI models over traditional bespoke transcription algorithms, typically based in optical character recognition (OCR) software, is their ability to generalize rapidly to new forms. Unlike past approaches (18,19), which were often tailored to specific vaccination card formats and required manual adaptation for new designs, generative, AI-based approaches as described in our approach are suitable for multiple layouts, with minimal need for bespoke software development nor long lead times for development. As we have shown, to achieve high-fidelity accuracy still requires engineering, development, and human curation, but the overall amount of time and expertise level is significantly reduced. Moreover, the speed at which generative AI models are improving is staggering; we expect for future models, less engineering and software development will be required to achieve even better results. This approach leveraging generative AI is more scalable and adaptable for use in diverse survey and healthcare settings, well beyond the narrow use case of childhood vaccination cards. However, while our dataset did include several card designs, further research is needed to evaluate how well these models generalize across an even broader range of formats, languages, and handwriting styles. Future work should explore fine-tuning on expanded datasets and testing AI performance across different countries and immunization programs to ensure that these models remain reliable and accurate in real-world deployments.

A key practical limitation—and a broader challenge to adoption and field implementation of this approach—is limited internet connectivity and the current inability of many multimodal foundation models to run on edge devices such as mobile phones or tablets. Additionally, the use of proprietary models (e.g., those developed by companies such as OpenAI (12) or Anthropic) may be restricted in settings where data must remain on-device or within national borders due to concerns around data sovereignty, privacy, and sensitivity—particularly for health-related information concerning children. Open-source foundation models, including small models that can run on peripheral devices and midsize models that can be deployed on in-country servers, may help mitigate these constraints. Emerging models like Phi-3.5 Vision (17) and LLaMA 3.2 offer multimodal capabilities and could represent promising alternatives. Importantly, deploying open-source models in controlled environments also enables researchers to probe and better understand underlying model biases, which is essential for ensuring safety, especially when handling sensitive health data. Evaluating how well these models perform on this task is a critical next step of this research.

This study serves as a proof of concept; additional research is needed to refine, validate, and operationalize AI-augmented transcription for vaccine coverage surveys. Moving toward real-world implementation will require further testing, optimization, and ethical considerations. A key next step is expanding validation across a broader dataset, assessing model performance across different vaccination card designs, languages, and handwriting styles to evaluate generalizability. Additionally, by integrating AI transcription into real-world immunization survey workflows, we can test data collection efficiency, cost, accuracy, and logistical constraints.

While this study focuses specifically on digitizing vaccination records, we envision broader implications for automated transcription and digitization of handwritten records across various domains. Paper-based records still exist widely either as data collections or in active use, including for example as vaccination registers in health facilities, medical records, survey forms, patient registries, and census documents. Multimodal AI models, with their generalizable utility, have the potential to integrate previously paper-based records into digital systems more efficiently than ever before. By automating transcription across diverse document types, they can reduce reliance on manual data entry, enhance data accessibility, and accelerate digitization efforts in various fields.

## Conclusion and Recommendation

AI-assisted transcription has the potential to significantly reduce the time and cost of large-scale immunization coverage surveys while preserving or improving data quality. Given these advantages, incorporating images of home-based records (HBRs) as a standard practice in vaccination coverage surveys would enable the integration of AI transcription tools into survey workflows. While some level of human review will remain necessary, confidence-based segmentation can ensure that manual verification is targeted efficiently, focusing only on cases with uncertain transcriptions. Furthermore, the generalizability of multimodal AI models extends beyond immunization, and our findings imply opportunities for digitizing handwritten records in many other domains.

Disclaimer: M.C.D.-H., and M.K. work for the World Health Organization (WHO). The authors alone are responsible for the views expressed in this publication and they do not necessarily represent the decisions, policy, or views of the WHO.

## Supplementary Information

### I. Limitations of Self-Assessment by the LLM

We explored whether prompting the large language model (LLM) to self-assess its own performance would yield useful diagnostic information. Specifically, we included the following instruction in the prompt: *“Self-assess transcription accuracy, sometimes called self-reflection. Finally, include your own assessment of handwriting clarity and visibility of date in the image. Your clarity assessment should range from 0 to 10, 0 being totally illegible, and 10 being perfectly clear*.*”* Despite this explicit request, the resulting self-assessments were inconsistent, often vague, and failed to correlate with objective measures of transcription accuracy or image quality. In some cases, the LLM reported high confidence and clarity scores for images that were poorly transcribed or highly ambiguous. Conversely, some accurate transcriptions were paired with low self-assessment scores. These findings suggest that, at least in our use case and dataset, LLM-based self-reflection did not provide a reliable or interpretable metric of performance. As such, we excluded these outputs from our final evaluation metrics.

